# Prediction of ROR1 and CAV1 expression from whole-exome sequencing of tumor tissue and plasma cfDNA to infer the efficacy of B013 plus nab-paclitaxel in triple-negative breast cancer

**DOI:** 10.64898/2026.09.30.26364376

**Authors:** Min Jiang, Dongming Zhang, Yi-Ching Hsueh, Yu Zhao, Shuhua Zhao, Qiangzu Zhang, Min Huang, Panjun Gao, Xiaohong Liu, Yuhan Liu, Qihuang Jin, Wenbin Dong, Jing Zhang, Ran Ran, Ruyan Zhang, Yanhui Fan, Juntao Xu, Guangming Tan, Chunli Zhang, Chunming Zhang, Gang Niu

## Abstract

Accurate prediction of response to targeted anticancer therapy frequently depends on the expression level of the drug target. However, transcriptomic profiling requires tumor tissue that is often unavailable, and circulating cell-free DNA (cfDNA) sequencing carries no expression information. Here, in an in silico companion study to a trial of ROR1 antibody plus nab-paclitaxel in triple-negative breast cancer (TNBC), we transformed whole-exome sequencing (WES) data into somatic activity profiles of signaling pathways (sAPSPs) using the previously published DAGM framework, and used them together with matched expression data to develop regression model sets predicting ROR1 and CAV1 expression. The model sets were applied to 10 trial participants (4 with tumor tissue WES, 6 with plasma cfDNA WES). Patients were classified as predicted-sensitive only when both ROR1 and CAV1 were predicted to be highly expressed. The prediction matched the observed best response in 9 of 10 patients; all three predicted-sensitive patients achieved a partial response. These exploratory findings support the feasibility of WES-based expression prediction for non-invasive response stratification and warrant prospective validation.

## Introduction

Triple-negative breast cancer (TNBC) accounts for the least favorable outcomes among breast cancer subtypes, largely because of tumor heterogeneity and a long-standing paucity of effective therapies other than chemotherapy^[1]^. Nab-paclitaxel is a frequently used backbone of first-line systemic therapy for advanced TNBC^[1]^. Receptor-tyrosine-kinase-like orphan receptor 1 (ROR1) is an oncofetal surface receptor that is re-expressed in breast cancer, is associated with aggressive behavior and adverse prognosis in TNBC, and is the target of several clinical-stage agents^[2–5]^. B013, a monoclonal antibody targeting ROR1, was evaluated in combination with nab-paclitaxel as first-line treatment for locally advanced or metastatic TNBC in a clinical trial (CTR20222396).

ROR1 is overexpressed in several human malignancies, is associated with aggressive disease in breast cancer, and supports tumor-cell growth in experimental models^[2, 3]^. Therapeutic targeting of ROR1 has been shown to be clinically feasible, including a phase I trial of the anti-ROR1 antibody cirmtuzumab in chronic lymphocytic leukemia and preclinical activity of the ROR1-targeting antibody-drug conjugate NBE-002^[4, 5]^. For the chemotherapy component, caveolae-mediated endocytosis is critical for cellular uptake of albumin and for response to albumin-bound chemotherapy^[6]^. Tumor caveolin-1 (CAV1) expression has been associated with the efficacy of nab-paclitaxel-containing regimens in breast cancer, both in a phase II trial of weekly nab-paclitaxel plus gemcitabine and in the neoadjuvant GeparSepto trial^[7, 8]^. Target or transporter expression is therefore a plausible determinant of who benefits from this regimen. However, measuring expression requires tumor RNA, which is frequently unavailable, not repeatable, or unrepresentative of the current molecular state of the disease.

Circulating tumor DNA analyzed by whole-exome sequencing (WES) offers a non-invasive genomic snapshot that can show high concordance with metastatic tumor tissue^[9, 10]^. Liquid-biopsy profiling has been extended to increasingly complex molecular features of tumors^[11]^. However, DNA-level data by themselves convey no information on gene expression. Computational approaches that interpret mutations beyond sequence fall into two broad categories. Methods such as xseq integrate paired somatic mutation and expression data from the same tumors within a hierarchical Bayesian framework, but are inapplicable to mutation-only specimens^[12]^. In contrast, the DAGM (Damage Assessment of Genomic Mutations) framework learns the cumulative impact of mutations on gene expression from reference data and projects an individual’s variants onto activity profiles of signaling pathways (APSPs), thereby enabling pathway-level functional readouts from mutation data alone^[13]^. In parallel, in silico clinical trials (ISCTs) can emulate patient cohorts and interventions by computational models. They are emerging as a complement to conventional trials, enabling candidate biomarkers, dosing strategies and eligibility hypotheses to be evaluated computationally before patients are exposed^[14, 15]^. In oncology, digital-twin and model-informed approaches have supported dose-response characterization and biomarker identification in early-phase trials^[14, 16–18]^. Most of these frameworks, however, depend on data modalities such as pharmacokinetic sampling that are not routinely collected for every participant. A virtual-trial framework built on WES alone would be directly compatible with standard genomic workups of tumor tissue or plasma.

Here we report an in silico companion study (NCT05833802) conducted in parallel with the trial of B013 plus nab-paclitaxel. In this study, we found that WES-derived sAPSPs can be used to predict the expression of ROR1 and CAV1 in TNBC, and further to infer which trial participants would respond to the combination.

## Materials and methods

### Patients and samples

The patients tested in this in silico companion study were from a clinical trial evaluating the efficacy and safety of B013 plus nab-paclitaxel as first-line therapy for locally advanced or metastatic TNBC (CTR20222396). Trial participants were treated, evaluated and followed according to the trial protocol, with efficacy assessed every eight weeks using RECIST v1.1. The best overall response was used for the present analysis, and duration of disease control (DDC), defined as the time from the first dose to the date of progressive disease per the overall response assessment, or to the date of the last imaging assessment for patients without progression, was recorded in days. The study was reviewed and approved by the Ethics Committee of Peking University Cancer Hospital & Institute (approval no. 2022YJZ109), and all participants provided written informed consent.

Model development used 40 patients with TNBC from a previously described local cohort^[13]^ for whom matched leukocyte WES, tumor tissue WES and tumor tissue RNA transcriptome data were available. The application cohort from the clinical trial (CTR20222396) comprised 11 participants with available WES data. One participant was excluded because efficacy could not be evaluated, leaving 10 participants for analysis. Among them, 4 had tumor tissue WES and 6 had plasma cfDNA WES collected before treatment.

### WES

Matched tumor tissue and peripheral blood were obtained from each patient. Peripheral blood (8 mL) was collected into Cell-Free DNA BCT tubes (Streck) and centrifuged at 1,600 x g for 10 min at 4°C. The separated plasma was further centrifuged at 16,000 x g for 10 min at 4°C to remove residual cells and platelets. Plasma cfDNA was extracted from approximately 4 mL of plasma using the QIAamp Circulating Nucleic Acid Kit (QIAGEN; 55114), and leukocyte genomic DNA (gDNA) was extracted from the remaining blood cell fraction using the Magnetic Universal Genomic DNA Kit (TIANGEN; DP705). Formalin-fixed paraffin-embedded (FFPE) sections (tumor cell content ≥ 20% on H&E staining) were deparaffinized with xylene, and DNA was extracted using the TIANamp FFPE DNA Kit (TIANGEN; DP331). gDNA was assessed by agarose gel electrophoresis and Qubit quantification, and cfDNA by the Qubit dsDNA HS Assay and an Agilent 2100 Bioanalyzer.

Libraries were constructed with the NEB Next Ultra II DNA Library Prep Kit (New England Biolabs; Z1723L). FFPE tumor DNA and leukocyte gDNA were sheared to 180-280 bp using a Covaris focused-ultrasonicator, whereas cfDNA was used without fragmentation to preserve its native size profile^[19]^. All libraries were captured with the same exome probe set (Agilent SureSelectXT Human All Exon V6; 5190-8864), quality-checked by Qubit, Bioanalyzer and qPCR, and sequenced on an Illumina NovaSeq 6000 platform (PE150) to mean depths of 350x (FFPE tumor), 794x (cfDNA) and 199x (leukocyte gDNA). Leukocyte gDNA served as the matched germline control for somatic variant calling and was also used to filter out clonal hematopoiesis-derived variants^[20]^. DNA extraction, library preparation and sequencing were performed by Novogene Co., Ltd. (Beijing, China).

### Variant calling and copy number analysis

Variant calling and copy number analysis were performed as previously described^[13]^. In brief, after removal of adapter-containing and low-quality reads, clean paired-end reads were aligned to the human reference genome using BWA. Duplicate reads were marked with Picard, followed by base quality score recalibration with GATK. Somatic variants in FFPE tumor DNA and cfDNA were called separately against the matched leukocyte gDNA. Single-nucleotide variants were detected using MuTect and small insertions and deletions using Strelka. Only calls passing default filters were retained. Variants were annotated with ANNOVAR and retained if they met all of the following criteria: (i) sequencing depth ≥ 20x with ≥ 5 variant-supporting reads and variant allele frequency (VAF) ≥ 5% in FFPE tumor DNA or ≥ 1% in cfDNA; (ii) depth ≥ 10x and no variant-supporting reads in the matched leukocyte gDNA; and (iii) population allele frequency ≤ 1% in the 1000 Genomes Project and gnomAD. Non-synonymous SNVs, splice-site variants and coding indels were used for subsequent analyses. Allele-specific somatic copy-number alterations, tumor cellularity and ploidy were estimated using Sequenza. Tumor mutational burden (TMB) and tumor fraction (TF) were calculated from the WES data as described previously^[9]^.

### Generation of sAPSPs

For each individual, somatic variants and copy-number alterations from tumor tissue or cfDNA WES were input into the previously published DAGM framework^[13]^. In brief, DAGM calculates the cumulative functional impact of genomic variants on global gene expression and maps these changes onto signaling pathways, generating a patient-specific sAPSP. The sAPSP served as the input features for expression prediction.

### Development of expression prediction model sets

The response variable was the target gene’s expression in tumor tissue, asinh-transformed TPM. The 40 TNBC patients were partitioned into a training set (n = 32) and a held-out test set (n = 8) by stratified sampling on the response variable (caret::createDataPartition). Feature engineering was performed on the training set only: an initial correlation-based filter (removing features with absolute Pearson correlation < 0.2 to the response), removal of highly collinear features (cutoff 0.85), and bootstrap aggregated feature selection (BAFS) consisting of 75% subsampling, correlation-filtered feature screening and stability counting over 100,000 repeated iterations, followed by top-feature selection and a final collinearity filter (cutoff 0.8).

Regression models were trained with the caret package in R 4.3.1. Of 118 configured regression algorithms, 112 ran successfully and were tuned by randomized parameter search with 10-fold cross-validation repeated 10 times. Models were then selected by cross-validation R^2^ ≥ 0.4 on the training set, followed by R^2^ ≥ 0.4 on the held-out test set.

### Statistical analysis

Correlations were assessed with Pearson or Spearman coefficients as indicated, and group comparisons with two-sided Wilcoxon rank-sum tests. P < 0.05 was considered statistically significant. Analyses were performed in R 4.3.1. Given the exploratory design and small sample sizes, no adjustment for multiple comparisons was applied, and results are reported descriptively.

## Results

### Study design and oversight

To investigate whether therapeutic efficacy can be predicted from tumor tissue WES, or even from plasma cfDNA WES, we conducted an in silico companion study in parallel with a clinical trial of B013 plus nab-paclitaxel as first-line therapy for locally advanced or metastatic TNBC (CTR20222396). First, expression-prediction model sets for ROR1 and CAV1 were established by machine learning using a previously described TNBC cohort^[13]^ as the modelling resource. The modelling resource comprised 40 patients with TNBC for whom tumor-tissue WES, matched white-blood-cell (WBC) DNA WES and tumor RNA-seq data were available. WBC WES provided the matched germline reference. Somatic variants and copy-number alterations (CNVs) from tumor WES were transformed by DAGM into sAPSPs, and tumor RNA-seq data provided the regression targets, ROR1 and CAV1 expression (Fig. 1A). Next, tumor tissue WES or plasma cfDNA WES were generated from TNBC patients enrolled in the clinical trial. The same DAGM step converted these data into tissue or cfDNA sAPSPs, which were fed into the established model sets to predict ROR1 and CAV1 expression and thereby infer patient sensitivity to B013 plus nab-paclitaxel (Fig. 1B).

**Figure 1.**
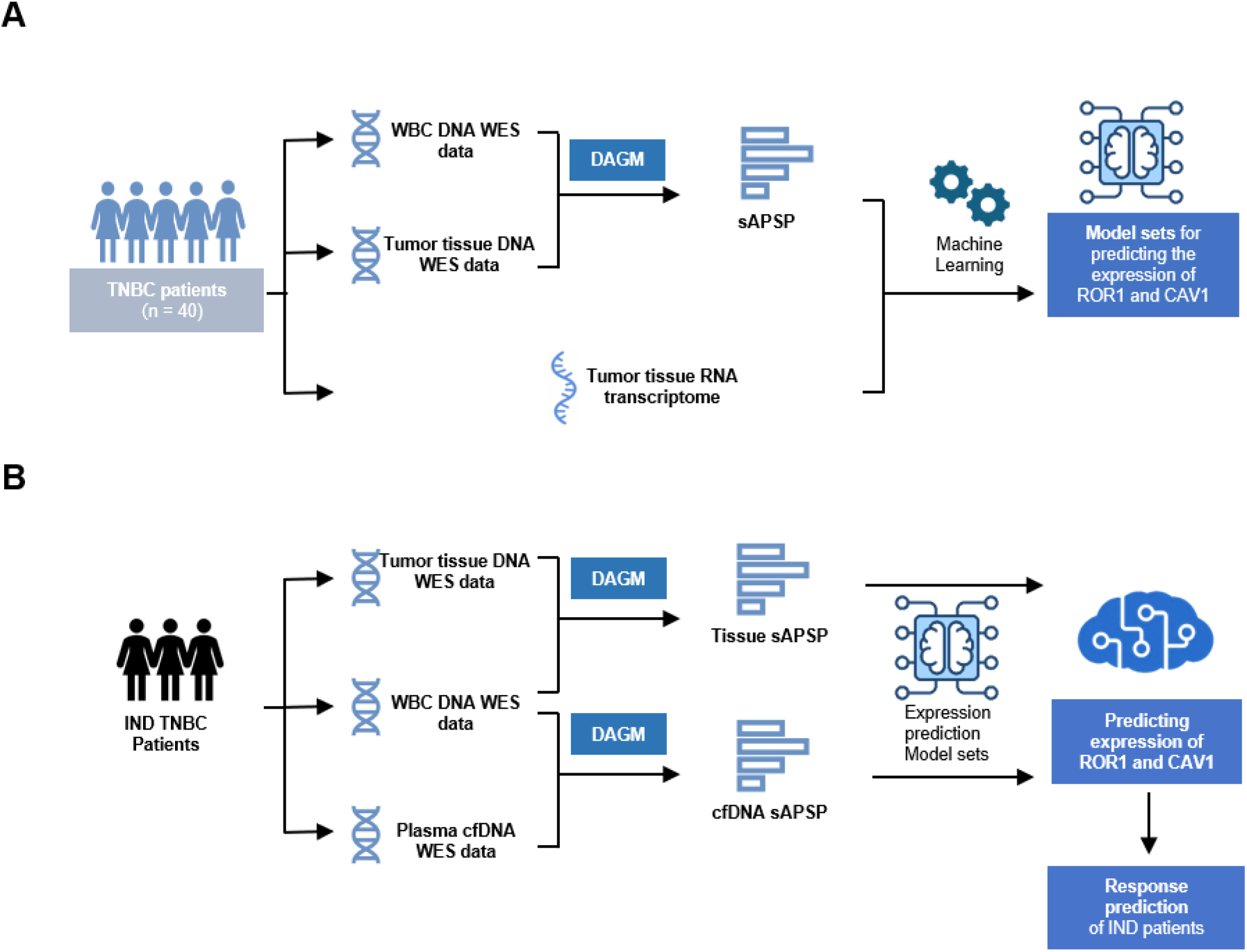
Study design and analytical workflow. (A) Workflow for establishing expression-prediction model sets for ROR1 and CAV1. Whole-exome sequencing (WES) data were transformed by the DAGM framework into somatic activity profiles of signaling pathways (sAPSPs), which served as predictors, and the matched tumor transcriptomes served as the regression targets. Machine learning was used to establish expression-prediction model sets for ROR1 and CAV1. (B) Application of the model sets to predict drug responses of clinical trial participants. WBC DNA WES together with either tumor tissue WES or plasma cfDNA WES was generated from each participant. DAGM was then used to generate tissue or cfDNA sAPSPs, which entered the model sets to predict the expression of ROR1 and CAV1, and in turn the response of participants to B013 plus nab-paclitaxel.

### Development of expression-prediction model sets for ROR1 and CAV1

We first asked whether a quantitative regression from pathway activity to gene expression could be learned at all from 40 patients and 3,074 candidate features. The 40 patients were partitioned into a training set (n = 32) and a held-out testing set (n = 8), stratified on the response variable and generated independently for each target. All feature engineering and model selection were performed within the training set, and the testing set was carried through passively (Fig. 2A). This feature-engineering procedure returned final feature sets of 5 features for ROR1 and 24 for CAV1, as detailed in Figure S1. A pool of 112 regression algorithms was then trained on the retained features with randomized hyper-parameter search under 10-fold cross-validation repeated 10 times. The resulting models were subjected to two-stage selection requiring a cross-validation R² ≥ 0.4 followed by a held-out R² ≥ 0.4 (Fig. 2A). Forty ROR1 models and 41 CAV1 models passed the cross-validation stage, of which 19 and 6, respectively, met the held-out criterion and constitute the final model sets (Fig. 2B). The strongest individual held-out performances were observed for the qrf model for ROR1 (test R² = 0.529) and the rlm model for CAV1 (test R ² = 0.612), and representative models of each set showed significant correlation between predicted and measured expression in held-out samples (ROR1: R = 0.78, P = 0.022; CAV1: R = 0.81, P = 0.014; Fig. 2C). Thus, expression-prediction model sets for ROR1 and CAV1 were established on the basis of WES-transformed sAPSPs.

**Figure 2.**
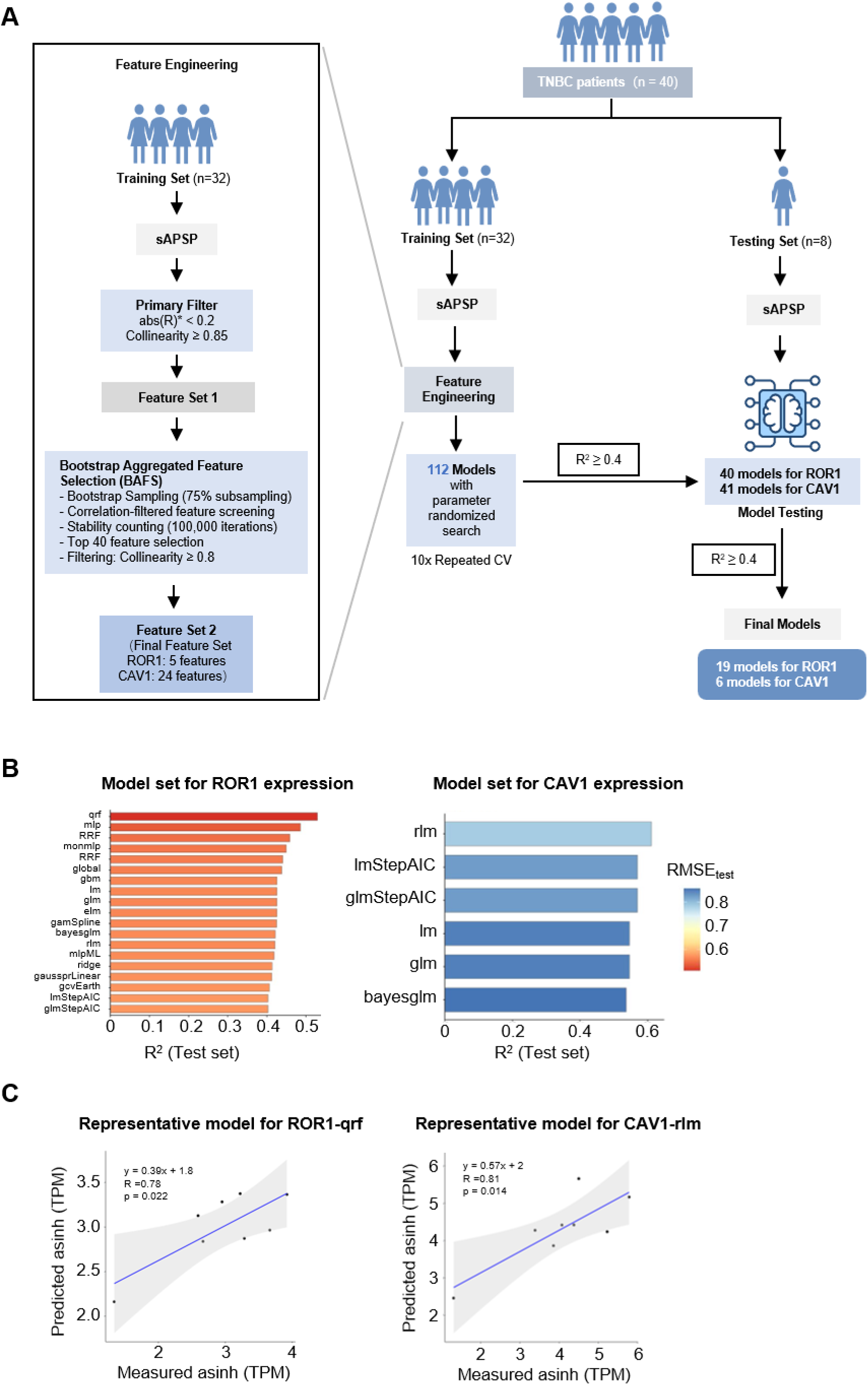
Development of expression-prediction model sets for ROR1 and CAV1. (A) Modelling workflow. The 40 TNBC patients were split into a training set (n = 32) and a testing set (n = 8). Feature engineering was performed, including a primary correlation filter, collinearity removal, and bootstrap-aggregated feature selection, yielding final feature sets of 5 (ROR1) and 24 (CAV1) sAPSP features. The features were used to train 112 regression algorithms with randomized hyper-parameter search under repeated 10-fold cross-validation. After filtering (R² ≥ 0.4), the remaining models were tested in held-out samples, and models with R² ≥ 0.4 were selected. (B) The final model sets of 19 ROR1 and 6 CAV1 models. Bar length indicates the coefficient of determination on the held-out testing set (R², test set) and bar color indicates the corresponding root-mean-square error (RMSE, test set). (C) Scatter plots of predicted versus measured expression in the held-out testing set (n = 8) for the representative models ROR1 (qrf) and CAV1 (rlm).

### Predicting therapeutic efficacy of B013 plus nab-paclitaxel in TNBC patients by the model sets based on tumor tissue WES

The model sets were first applied to the four trial participants with tumor tissue WES, yielding per-patient distributions of predicted ROR1 and CAV1 expression across each model set (Fig. 3A). Each gene was classified as predicted-high or predicted-low relative to the cohort consensus, and a participant was inferred to be sensitive only when both genes were predicted high. The inferred sensitivity matched the observed best overall response in three of the four participants (Fig. 3B). P02 was called ROR1 High and CAV1 High, was predicted sensitive, and achieved a partial response (PR) with a DDC of 273 days. P01 (ROR1 High, CAV1 Low) and P03 (ROR1 Low, CAV1 Low) were predicted non-sensitive and both had stable disease (SD), with DDC of 156 days and more than 273 days, respectively. P04 (ROR1 Low, CAV1 High) was predicted non-sensitive but achieved a PR with a DDC of 161 days. Although the small sample size precludes informative inferential statistics, this indicates the feasibility of predicting therapeutic efficacy from WES data alone.

**Figure 3.**
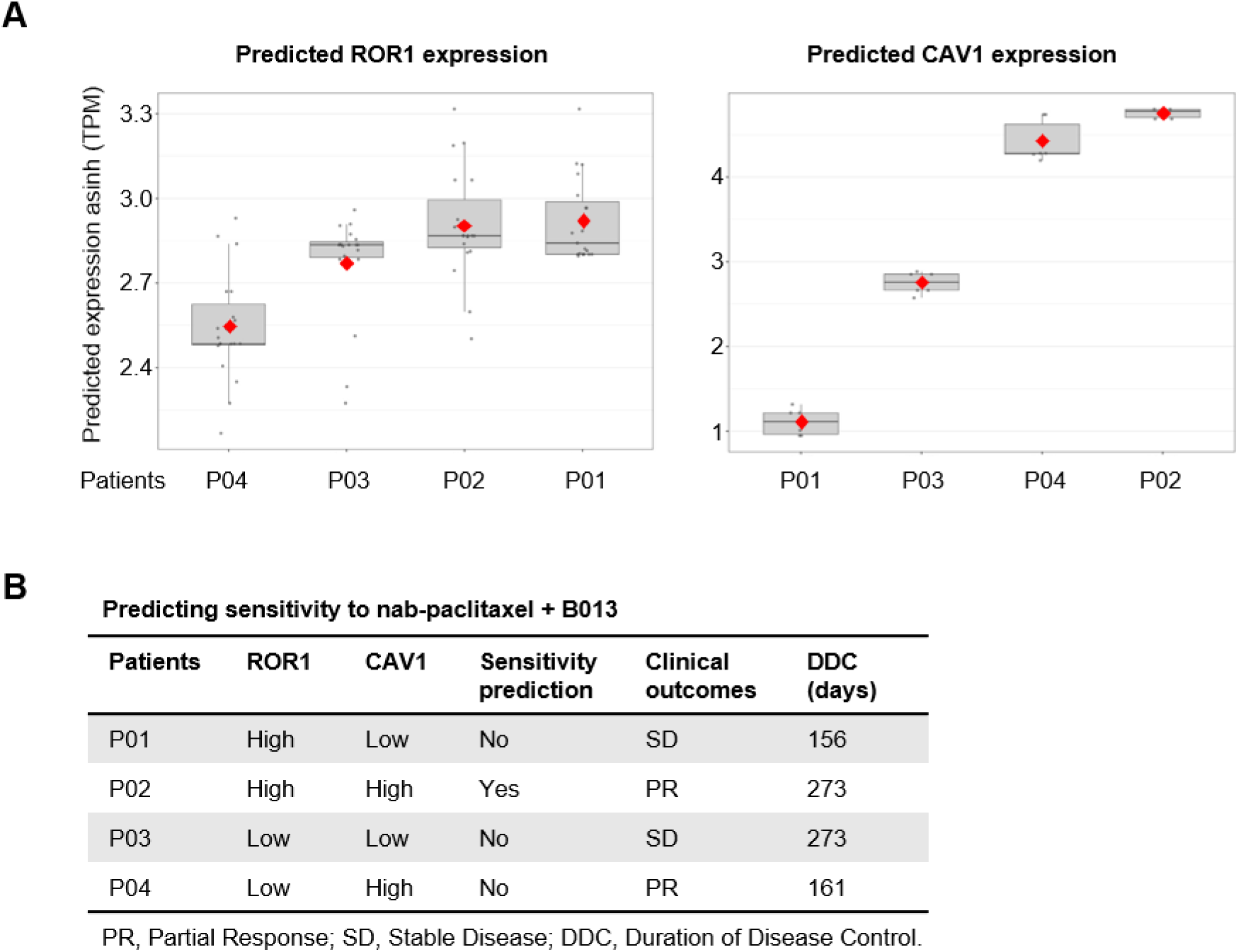
Predicting therapeutic efficacy of B013 plus nab-paclitaxel in TNBC patients by the model sets based on tumor tissue WES. (A) The expression of ROR1 and CAV1 was predicted by the expression-prediction model sets based on tumor tissue WES. Boxplots summarize the distribution across the model set (diamond, mean). (B) Prediction of therapeutic efficacy of B013 plus nab-paclitaxel. ROR1 and CAV1 are called High or Low relative to the cohort consensus. A patient is predicted sensitive only when both are High. Clinical outcome is the best overall response by RECIST v1.1. PR, partial response; SD, stable disease; DDC, duration of disease control. “>” indicates that disease control was ongoing at data cut-off.

### Predicting therapeutic efficacy of B013 plus nab-paclitaxel in TNBC patients by the model sets based on plasma cfDNA WES

We next asked whether the same inference could be achieved non-invasively from plasma cfDNA WES. All 5 ROR1 and 24 CAV1 model features were detectable in cfDNA WES from the trial participants (Fig. S1). Feature values from cfDNA and tissue samples fell within the range of the training data (training: −4.17 to 4.47; trial cohort: −4.32 to 4.29), and trial samples did not form isolated clusters in the feature space. In addition, the tumor-derived DNA fraction in plasma cfDNA approached or exceeded 20% (Fig. 4A). It has been reported that the concordance of clonal somatic mutations and CNVs between tumor tissue and patient-matched ctDNA with a tumor fraction above 10% was 88% and 80%, respectively^[9]^. These data suggest that the model sets can be applied to cfDNA-derived inputs to predict the expression of ROR1 and CAV1.

**Figure 4.**
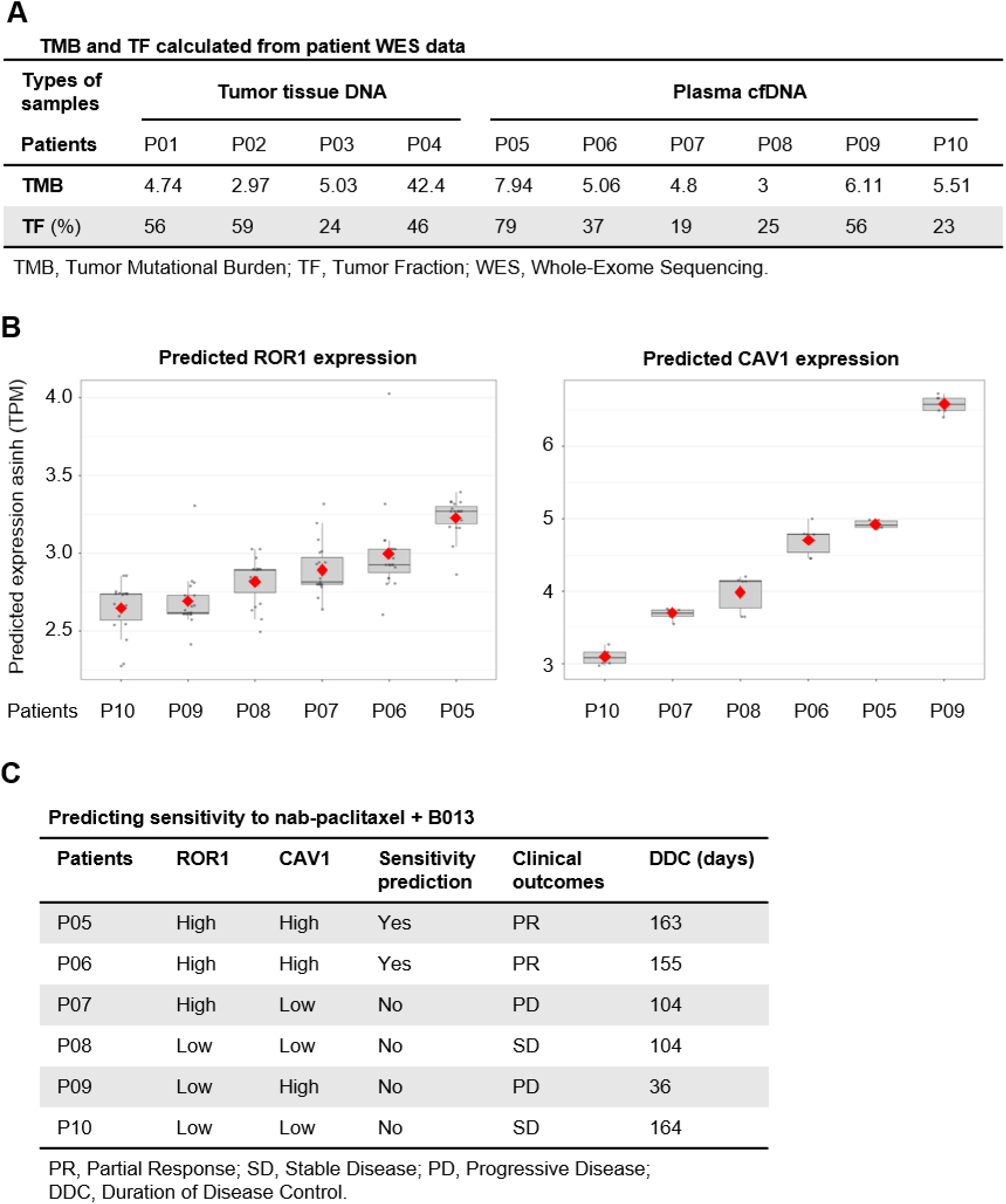
Predicting therapeutic efficacy of B013 plus nab-paclitaxel in TNBC patients by the model sets based on plasma cfDNA WES. (A) Tumor mutational burden (TMB) and tumor fraction (TF) calculated from WES data of all ten trial participants. TF denotes tumor cell content estimated from tumor tissue WES for P01–P04 and the circulating tumor-derived DNA fraction estimated from plasma cfDNA WES for P05–P10. (B) The expression of ROR1 and CAV1 was predicted by the expression-prediction model sets based on plasma cfDNA WES. Boxplots summarize the distribution across the model set (diamond, mean). (C) Prediction of therapeutic efficacy of B013 plus nab-paclitaxel. ROR1 and CAV1 are called High or Low relative to the cohort consensus. A patient is predicted sensitive only when both are High. Clinical outcome is the best overall response by RECIST v1.1. PR, partial response; SD, stable disease; PD, progressive disease; DDC, duration of disease control. “>” indicates that disease control was ongoing at data cut-off.

We then repeated the analysis using pre-treatment plasma cfDNA WES from the six trial patients P05–P10, with prediction, consensus scoring and the sensitivity rule identical to those in the tissue arm (Fig. 4B). All six patients were concordant with the observed outcome (Fig. 4C). P05 and P06, both called ROR1 High and CAV1 High, were predicted sensitive and both achieved a PR, with DDC of 163 and 155 days, respectively. P07 (ROR1 High, CAV1 Low) and P09 (ROR1 Low, CAV1 High) were predicted non-sensitive and both had progressive disease (PD), with DDC of 104 and 36 days, respectively. P08 and P10, both called Low for both targets, were predicted non-sensitive and both had SD, with DDC of more than 104 days and 164 days, respectively.

Considered together with the tissue arm, the rule agreed with the best overall response in 9 of 10 patients, and all three patients predicted sensitive across both specimen types achieved a PR. Given the small sample size, statistically significant results could hardly be expected. Nevertheless, these results demonstrate the feasibility of predicting therapeutic efficacy from non-invasive plasma cfDNA WES alone.

## Discussion

In this in silico companion study conducted alongside a trial of B013 plus nab-paclitaxel in TNBC, we show that WES-derived sAPSPs can be used to train regression model sets that predict ROR1 and CAV1 expression. Ranking the predicted expression of ROR1 and CAV1 then allowed us to infer TNBC patient response to B013 plus nab-paclitaxel, with all three predicted-sensitive participants achieving a PR. We thus established a method for predicting drug response based solely on WES data, which can even be obtained from plasma cfDNA, a non-invasive source.

Our approach builds on the DAGM framework, which translates mutation data into pathway-activity profiles^[13]^. Whereas DAGM was originally applied to germline-based risk assessment in HER2-negative breast cancer, we applied it to somatic WES from tumor tissue and cfDNA and extended the readout from pathway activity to gene-level expression. The mutation-to-expression inference problem has previously been addressed by xseq using paired mutation – expression data within the same tumors^[12]^. Our strategy is complementary in that it requires mutation data only, at the cost of reduced per-patient certainty. The detectability of tumor-derived variants in cfDNA WES and their concordance with metastatic tissue^[9–11]^ are consistent with our observation that all model features were measurable in cfDNA. Our work also connects to digital-twin and in silico trial methodologies that have supported dose-response characterization and biomarker proposals in early-phase oncology^[14, 16–18]^. Unlike imaging- or pharmacology-based twins, the present twins are built solely from WES.

Expression-level information has previously been recovered from plasma through the physical state of the cfDNA molecule rather than through its sequence. Nucleosome-depletion patterns at transcription start sites in whole-genome plasma sequencing can distinguish expressed from silent genes^[21]^, and promoter fragmentation entropy measured by targeted deep sequencing (EPIC-seq) yields quantitative estimates of individual gene expression in plasma^[22]^. These approaches read the chromatin state carried by circulating fragments and have been benchmarked directly against matched RNA. The present study takes a different route, in which the somatic variant and copy-number content of the exome is transformed into sAPSPs that are then used to model tumor gene expression. The two methods are complementary and may be integrated to produce better models for predicting gene expression in tumors.

The five ROR1 model features are pathway-activity measures related to purine nucleotide and amino-acid metabolism, and the 24 CAV1 features span inflammatory signaling and lipid and carbohydrate metabolism (Fig. S1). Because features were selected to optimize expression prediction rather than outcome prediction, these pathways should be interpreted as statistical correlates of the expression programs of ROR1 and CAV1, not as mechanistic drivers.

The rationale connecting predicted expression to drug response derives from prior experimental and clinical literature. ROR1 expression is a prerequisite for engagement by anti-ROR1 antibodies^[2–5]^, and CAV1-mediated endocytosis facilitates albumin-bound drug uptake^[6–8]^. Our data are consistent with these links but do not test them. Notably, one participant inferred non-sensitive (P04; predicted ROR1-low) achieved a PR, indicating that response can occur without a both-high prediction.

ROR1-directed therapy is an active area of clinical development. The ROR1-targeting antibody-drug conjugate zilovertamab vedotin showed clinical proof of concept, with objective responses in mantle cell lymphoma and diffuse large B-cell lymphoma, in a first-in-human phase 1 trial^[23]^. In breast cancer, the anti-ROR1 antibody zilovertamab combined with paclitaxel was well tolerated and associated with partial responses in a phase 1b study of HER2-negative disease^[24]^. Notably, that first-in-human trial enrolled patients unselected for tumor ROR1 expression^[23]^, so tissue-based assessment of the target was not used to guide treatment allocation. Genomic inference of target expression, as explored here, addresses this unmet need.

If validated prospectively with predefined thresholds, cfDNA-based expression inference could provide a non-invasive, repeatable, pre-treatment stratification tool to enrich for likely responders in trials of ROR1-targeted combination therapy, and could in principle be extended to longitudinal monitoring. Serial ctDNA assessment is already clinically informative in breast cancer, showing that post-neoadjuvant ctDNA positivity identifies patients with TNBC at high risk of recurrence^[25]^. These results indicate that repeated plasma sampling is feasible and interpretable in this disease. Expression inference from serial cfDNA could, in principle, add a functional layer to such monitoring. More broadly, this workflow illustrates how companion in silico analyses can be embedded into early-phase drug development to refine eligibility hypotheses before committing patients. These implications remain investigational.

There are several limitations to this study. The application cohort is small, with only 10 evaluable participants, and at this size formal statistical confirmation is not achievable. Unfortunately, we could not test additional participants because the trial had been terminated. The predicted expression of ROR1 and CAV1 in trial participants was not confirmed by direct measurement, so the correctness of the inferred expression itself remains untested in these patients. In the future, better models could be trained using WES from tissue DNA and plasma cfDNA together with matched expression data in a larger, independent TNBC cohort. It would be promising to extend the approach to longitudinal cfDNA sampling for dynamic response monitoring.

In conclusion, WES-derived sAPSP model sets can predict ROR1 and CAV1 expression in TNBC, and further infer patient response to B013 plus nab-paclitaxel. Predictions were concordant with observed responses in 9 of 10 evaluable trial participants, with complete concordance for the non-invasive cfDNA approach in six participants. These findings support further development of WES-based expression inference as a research tool for response stratification.

## Data Availability

All data produced in the present study are available upon reasonable request to the authors

## Acknowledgements

This study is funded by Beijing Natural Science Foundation (L232032) and Beijing Municipal Science & Technology Commission (No. Z231100004623006).

## Disclosing the use of artificial intelligence

An AI-assisted language tool (Kimi, China) was applied only for languish polishing. All study design, conceptualization, methodology, visualization, interpretation, and scientific writing were performed by the authors.

## Supplemental figure

**Figure S1.**
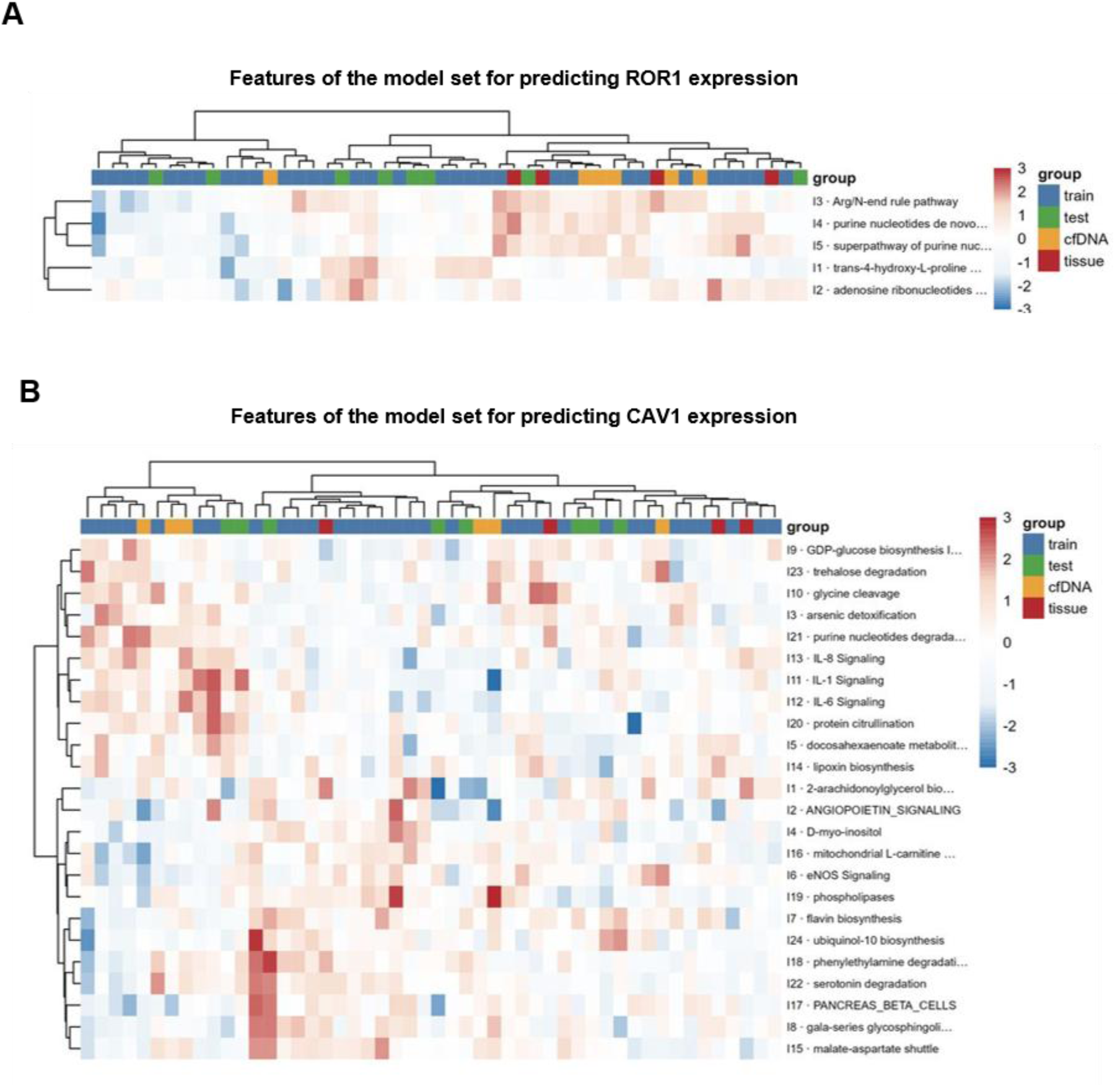
Model features are detectable in cfDNA WES. Heatmaps show the model-set input features, 5 for ROR1 (A), 24 for CAV1 (B). train, the samples for model training; test, the samples for model testing; cfDNA, IND participant plasma cfDNA samples; tissue, IND participant tissue samples.

